# Physiological foundation modeling for subclinical disease assessment: a prospective pilot

**DOI:** 10.1101/2025.08.20.25334090

**Authors:** William Yuan, Shiwei Xu, Sara Dionisi, Katharine von Herrmann, Kate Frederick-Dyer, Joyce Cheung-Flynn, Caleb Kennedy, Charles R Flynn, Scott Lipnick

## Abstract

**Objective:** Clinical studies struggle to locate the right patients, in part because many remain undiagnosed or lack relevant labels. We develop and prospectively test a physiology-based patient representation (‘Bioprofile’) to evaluate whether it can reduce the number of individuals who must be screened and enable more precise targeting of candidates for steatotic liver disease studies.

**Materials and Methods:** We trained Bioprofile patient representations from routinely collected health data and fine-tuned against multiple endpoints. Bioprofiles were trained using 1 million subjects from the UK Biobank and other cohorts. The trained model was applied to 45,484 research subjects at Vanderbilt University Medical Center. Based on Bioprofile nominations, 31 subjects were recruited for a prospective validation study. The primary outcome measure was proton density fat fraction (PDFF), an imaging-based metric of steatosis severity.

**Results:** Bioprofile models achieved a Spearman coefficient of 0.65 against PDFF, outperforming existing foundation models (ρ = 0.361) and clinical risk scores (ρ = 0.522-0.542) in retrospective validation. Simulations found that Bioprofiles required half as many subjects needed to screen compared to existing methods depending on task. Bioprofiles were further validated against 3 global cohort studies. Bioprofile predictions aligned strongly with prospective study data (ρ = 0.740).

**Discussion and Conclusion:** AI-based profiling of patient physiology can reveal individuals who have subclinical signatures of disease. If implemented widely, this approach can identify unknown human subjects, reduce screening failures, and improve trial quality. Bioprofiles may have additional utility in decision support applications in precision medicine and AI-augmented healthcare.

## Introduction

Metabolic dysfunction-associated steatotic liver disease (MASLD; formerly non-alcoholic fatty liver disease, NAFLD^1^) is highly prevalent yet remains markedly underdiagnosed in clinical practice^2^. The condition, defined by liver fat accumulation (hepatic steatosis) in the presence of metabolic risk factors, affects over a quarter of adults worldwide^3^. However, many individuals with MASLD are never diagnosed, owing to its often subclinical course and the limited use of definitive diagnostics in routine care^4–6^. This poses a major challenge for patient recruitment in clinical studies, as potentially eligible participants may be missed. Even when MASLD is suspected, confirming steatosis severity typically requires imaging or biopsy, introducing logistical or financial hurdles in human trials^7,8^. MRI-based proton density fat fraction (PDFF) measurements are considered a gold-standard for noninvasive quantification of liver fat and are commonly used as an inclusion criterion for clinical trials. Changes in PDFF are also used as an endpoint of treatment efficacy for steatosis reduction^9–11^ However, MRI is expensive, time-intensive, and not accessible at scale^12,13,14^.

There is a clear need for steatosis metrics that are both non-invasive and scalable to flag high-risk patients. Simple clinical scores^15,16^ estimate MASLD probability from routine labs and demographics but offer only moderate accuracy that is insufficient to precisely quantify liver fat or guide trial enrollment. One recent study^17^ developed an artificial intelligence (AI) algorithm for identifying subjects based on electronic health records (EHRs) at particularly high risk and hypothesized that such an algorithm could support earlier screening and even facilitate recruitment for trials. However, this algorithm was only externally validated on retrospective real-world data where true disease labels were unavailable. Furthermore, human studies, particularly those focused on discovery efforts, may aim to identify subjects at various stages of disease, rather than just one. More broadly, foundation models, which utilize broad pre-training to enable prediction of multiple down-stream endpoints, have been applied to a wide set of healthcare applications, but liver specific applications and real-world validation have not been performed to our knowledge^18^.

In this work, we introduce and validate Bioprofiles, a foundation model of human physiology trained on large, diverse cohorts, first applied in a cardiometabolic context to detect patterns linked to liver-fat accumulation. By integrating clinically accessible data (demographics, medical history, laboratory results, and lifestyle factors), Bioprofiles can estimate an individual’s liver fat distribution without additional tests or imaging. Bioprofiles offer a non-invasive, easy-to-use tool to nominate likely MASLD patients before any intervention or imaging is done, thereby broadening the pool of trial-eligible participants. Outside of a trial setting, steatosis identification can contribute to qualifying patients for glucagon⍰like peptide⍰1 receptor agonists (GLP1 RAs)^19^,enabling earlier therapeutic engagement. In this manuscript, we evaluate Bioprofiles in a prospective pilot for MASLD trial recruitment.

## Methods

### Data sources

Deidentified data from participants in the UK Biobank^20^ (n = 500k), NHANES^21^ (n = 90k) and an internal set of hospital-based cohort data (n = 460k) were included in this study. The UK Biobank was subdivided into pretraining (n = 450k) and fine-tuning (n = 43k) sets based on availability of liver imaging data. Summary statistics from a set of 45,484 subjects from Vanderbilt University Medical Center (VUMC) were also utilized for calibration.

### Step 1: Model training

The primary endpoint considered was PDFF, measured using magnetic resonance imaging (MRI), which measures the relative degree of contributing signal from fat versus water and is expressed as a percentage value^22^. Clinically-accessible data elements including demographics, medical history, lab results, and lifestyle factors were extracted and manually harmonized across datasets. Factors were chosen based on what would plausibly be available or accessible during a primary care encounter. ReMasker^23^, a masked autoencoder originally designed for tabular imputation, was used to generate patient-level embeddings (Supplementary Methods, Supplementary Figure 3). Because PDFF is known to exhibit heteroscedasticity^24^ we utilized CARD,^25^ a method utilizing diffusion models for tabular regression and classification tasks that generates fuller conditional distributions of predicted PDFF, rather than just an estimate of mean and variance, enabling more sensitive estimates of predictive uncertainty. The CARD implementation from lightning-uq-box 0.2.0 was used. Hyperparameters for both ReMasker and CARD were optimized using Optuna 3.0.4 (Supplementary Table 6-7). All model building was performed using Torch 2.6.0 and Python 3.10 using an NVIDIA A10G GPU (Supplementary Figure 1A). The fine-tuning set was itself split into non-overlapping training, test, and validation sets (65:10:25). To ensure model performance was properly evaluated over the full spectrum of PDFF, we resampled the test set to a uniform distribution over PDFF bins.

To benchmark performance, we compared Bioprofile predictions to those generated by CLMBR, a recent foundation model that utilizes longitudinal electronic health records^26,27^, the Framingham Steatosis Index (FSI)^15^, and Hepatic Steatosis Index (HSI)^16^. We also employed two technical benchmarks: a gradient boosted regressor trained over the fine-tuning set, with access to i) body mass index (BMI), alanine aminotransferase (ALT), and aspartate aminotransferase (AST) levels, and ii) all Bioprofile input features. Bioprofile performance was also evaluated relative to the full technical benchmark on a series of non-proton density fat fraction (PDFF) tasks (Supplementary Table 1).

### Step 2: Targeted recruitment task

The first recruitment task we considered was to identify subjects with PDFF > 10%. This corresponds to a common inclusion criterion for interventional trials, where sponsors are interested in ensuring that recruited subjects have a sufficient amount of steatosis^28,29^. In simulated studies and during the VUMC recruitment pilot, we aimed to minimize the number of subjects screened to recruit 12 subjects with PDFF > 10%. The specific recruitment targets for the study were derived from sample size (Supplement) and logistical study considerations.

### Step 3: Even recruitment task

The second recruitment task we considered was to identify a uniform number of subjects across PDFF, specifically PDFF between 0% and 5%, between 5% and 15%, and above 15%, corresponding to healthy, mild, and moderate degrees of steatosis^30^. These correspond to a discovery-related task where a sponsor is interested in obtaining a sufficient number of subjects across a transition between relative health and disease, for the purpose of developing or validating a diagnostic or biomarker^31^. In simulated studies and during the VUMC recruitment pilot, we aimed to minimize the number of subjects screened to recruit at least 9 subjects across all 3 bins.

### Step 4: Cohort nomination algorithm

We developed an algorithm (Portfolio Selection, Supplementary Figure 1B) to nominate subjects for recruitment based on CARD-generated conditional probability distributions over predicted PDFF. We utilized linear programming to nominate subcohorts of subjects that minimized the Wasserstein distance between i) the collective conditional PDFF distribution of the nominated subcohort and ii) the goal distribution (all subjects with PDFF > 10% or even distribution of PDFF over 3 bins). Nominated subjects would be recruited and measured, and a new goal distribution defined if needed. Constraints such as recruitment batch size, latency between cohort identification, and measurement were simulated. Notably, for the VUMC study, we were able to embed the targeted recruitment task within the even recruitment task, identifying a single recruitment policy that simultaneously met both criteria (Supplementary Figure 1C). The Portfolio Selection algorithm is described in the Supplement and was trained using PuLP 2.7.0 and the PULP_CBC_CMD solver.

### Step 5: Study recruitment simulations

To evaluate the difficulty of obtaining cohorts with desired distributions, we simulated the ability of various models to guide subject recruitment studies across both tasks (targeted and even recruitment). These experiments were performed using the test set without label resampling. Furthermore, we evaluated two methods for model-guided subject recruitment: standard sampling, and Portfolio Selection. For standard sampling in targeted recruitment, subjects were recruited in rank order based on their model-assessed probability of having PDFF > 10%. For standard sampling in even recruitment, subjects were recruited in batches of 5 based on rank order of model-assessed probability of membership in each of the three bins, with recruitment shifting across bins as recruitment goals are reached (adaptive sampling).

### Subject screening and pre-recruitment

To operationalize the model-guided recruitment, we first utilized the Bioprofile and Portfolio Optimization algorithms to define a recruitment policy that would minimize the number of required recruits to achieve the goal distribution. This policy consisted of a set of several model-defined subpopulations to be recruited in a certain ratio. We used knowledge distillation with Bioprofile/Portfolio Optimization labels (teacher) to train a set of logistic regression classifiers (student) that were deployed at VUMC. We found that the student model had comparable performance to the teacher in terms of projected study performance (Supplementary Table 4). VUMC health records in the Epic data warehouse were eligible if they were between the ages of 21 and 70. Exclusion criteria for the fine-tuning cohort and VUMC recruitment cohort included pregnancy or breast-feeding, presence of implanted cardiac defibrillator or pacemaker, presence of cancer and/or ongoing cancer treatment, presence of HIV, history of alcohol consumption exceeding 14 drinks per week for men and greater than seven per week for women, illicit drug use, treatment with any investigational drug in the one month preceding study, mental conditions rendering an inability to understand the nature, scope and possible consequences of the study, inability to comply with the protocol (Supplementary Information). We also excluded subjects with existing diagnoses of MASLD from the VUMC recruitment cohort to evaluate the utility of our models on identifying undiagnosed MASLD. The study was approved by the Vanderbilt IRB (230833).

### Subject recruitment and sampling

A recruitment cohort of 45,484 subjects was identified in the VUMC clinical data warehouse based on inclusion, exclusion, data density, and subject accessibility requirements (Supplementary Information). Bioprofile student models were deployed on data present in their medical record (Pre-visit EHR Extract) and used to nominate subjects for recruitment, but a full physical exam and lab series was performed at the study visit (Study Visit). Data on all cohorts are provided in Table 1. After the study visit, Bioprofile teacher models were deployed on deidentified pre-visit and study visit data. A CONSORT diagram of participant recruitment is provided in Supplementary Figure 2. Participants were compensated for their participation in the study. No adverse reactions were reported as part of participation.

**Table 1:** Characteristics of the pretraining, fine-tuning, and recruited cohorts. All values are means (95% confidence interval) unless otherwise indicated.

|  | Pretraining Cohort<br>(1M) |  | UKB Imaging Cohort (n=43k) |  |  | VUMC Cohort (Pre-Visit<br>EHR Extract)<br>(n=29) (2 ineligible for MRI) |  |  | VUMC Cohort (Study Visit)<br>(n=29) (2 ineligible for MRI) |  |  |
| --- | --- | --- | --- | --- | --- | --- | --- | --- | --- | --- | --- |
|  | Metric | Fraction<br>with Data | PDFF <<br>5% (32k) | 5% <<br>PDFF <<br>15%<br>(9.1k) | PDFF ><br>15%<br>(2.1k) | PDFF<br>< 5%<br>(10) | 5% <<br>PDFF<br>< 15%<br>(9) | PDFF<br>> 15%<br>(9) | PDFF<br>< 5%<br>(11) | 5% <<br>PDFF<br>< 15%<br>(9) | PDFF<br>> 15%<br>(9) |
| Age (years) | 49.61<br>(49.58 -<br>49.63) | 1 | 55.01<br>(54.93 -<br>55.09) | 55.47<br>(55.32 -<br>55.63) | 54.1<br>(53.85 -<br>54.42) | 50.06<br>(40.42<br>-<br>58.54) | 41.45<br>(33.63<br>-<br>48.51) | 43.28<br>(37.77<br>-<br>49.12) | 50.55<br>(40.18<br>- 60) | 41.49<br>(34.5 -<br>50.13) | 43.55<br>(36.19<br>-<br>50.23) |
| Sex (fraction<br>male) | 0.486<br>(0.485 -<br>0.487) | 1 | 0.439<br>(0.434 -<br>0.443) | 0.606<br>(0.597 -<br>0.616) | 0.576<br>(0.556 -<br>0.597) | 0.618<br>(0.25 -<br>0.875) | 0.533<br>(0.163<br>-<br>0.777) | 0.644<br>(0.333<br>-<br>0.888) | 0.618<br>(0.25 -<br>0.875) | 0.533<br>(0.163<br>-<br>0.777) | 0.644<br>(0.333<br>-<br>0.888) |
| BMI (kg/m2) | 25.29<br>(25.28 -<br>25.3) | 0.992 | 25.65<br>(25.61 -<br>25.69) | 28.84<br>(28.76 -<br>28.93) | 30.53<br>(30.36 -<br>30.77) | 23.39<br>(21.57<br>-<br>26.34) | 38.29<br>(33.91<br>-<br>43.88) | 36.42<br>(32.78<br>-<br>40.98) | 23.52<br>(21.12<br>-<br>26.31) | 37.14<br>(32.41<br>-<br>41.44) | 37.47<br>(33.05<br>-<br>40.95) |
| Triglycerides | 1.55<br>(1.54 - | 0.955 | 1.47<br>(1.46 - | 2.04<br>(2.07 - | 2.29<br>(2.24 - | 0.683<br>(0.484 | 1.48<br>(1.27 - | 2.67<br>(1.69 - | 0.698<br>(0.52 - | 1.47<br>(1.28 - | 2.94<br>(2.1 - |
|  | 1.55) |  | 1.48) | 2.12) | 2.35) | - 0.88) | 1.67) | 3.92) | 0.863) | 1.69) | 3.81) |
| Serum ALT<br>(U/L) | 23.11<br>(23.08 -<br>23.14) | 0.923 | 20.7<br>(20.58 -<br>20.83) | 28.12<br>(27.86 -<br>28.41) | 35.68<br>(34.82 -<br>36.79) | 21.69<br>(15.93<br>-<br>28.32) | 49.16<br>(27.22<br>-<br>73.63) | 71.81<br>(47.68<br>-<br>96.06) | 21.66<br>(15.61<br>-<br>26.57) | 48.37<br>(27.39<br>-<br>74.78) | 78.8<br>(56.93<br>-<br>101.7) |
| Serum AST<br>(U/L) | 24.95<br>(24.93 -<br>24.98) | 0.801 | 25<br>(24.91 -<br>25.13) | 27.41<br>(27.16 -<br>27.67) | 30.6<br>(30.1 -<br>31.16) | 27.27<br>(21.18<br>-<br>34.45) | 31.86<br>(24.94<br>-<br>42.19) | 42.29<br>(30.15<br>-<br>54.65) | 27.56<br>(21.93<br>-<br>33.32) | 31.53<br>(25.05<br>-<br>39.51) | 47.08<br>(33.88<br>-<br>64.11) |
| Diabetes<br>Status | 0.043<br>(0.043 -<br>0.044) | 0.767 | 0.0168<br>(0.0154 -<br>0.0182) | 0.0511<br>(0.0475 -<br>0.0561) | 0.0704<br>(0.060 -<br>0.082) | 0 (0 -<br>0) | 0.57<br>(0.222<br>-<br>0.888) | 0.755<br>(0.444<br>-<br>1) | 0 (0 -<br>0) | 0.539<br>(0.222<br>-<br>0.836) | 0.793<br>(0.555<br>-<br>1) |
| PDFF | N/A | 0 | 2.60<br>(2.59 -<br>2.61) | 8.36<br>(8.31 -<br>8.41) | 21.0<br>(20.7 -<br>21.2) | N/A | N/A | N/A | 2.10<br>(1.73 -<br>2.46) | 9.90<br>(7.59 -<br>11.7) | 30.1<br>(24.9 -<br>35.0) |

The MyHealth@Vanderbilt platform was used to consent and engage nominated subjects. Recruited subjects were invited to participate in a study visit consisting of body measurements, venipuncture, blood analyses, and liver magnetic resonance imaging with the purpose of measuring PDFF.

### Subject schedule of study events

31 subjects who met inclusion and exclusion criteria were invited to the Vanderbilt Clinical Research Center after an overnight fast. A brief physical examination including measures of waist, hip, and chest circumference along with blood pressure were taken prior to a blood draw for a lipid panel, CBC, CMP and measurement of HbA1c. 2 subjects were found to be incompatible with MRI screening during the study visit. 29 subjects underwent MRI evaluation of the upper abdomen using the mDixon protocol. Results of estimated percentage of liver fat by MR method and Bioprofile-predicted liver fat content were correlated.

MRI examination of liver was done with a 3T MR7700 MR system (Philips Healthcare, Netherlands) with a quadrature body transmit coil used in combination with a 32-channel anterior receive coil array and 16-channel posterior receive coil array. Anatomic imaging of entire upper abdomen was performed with axial, free-breathing single-shot Turbo Spin Echo (TSE) sequence. A commercially available version of mDIXON sequence package was used to acquire fat and water images of entire liver. Following were the mDIXON Quant imaging parameters: 3D T1-FFE sequence, FOR=400/350/231mm, acq. vox.dim.=3/3/6mm (over contiguous slices with 3mm overlap), 6-echoes: TE1/delta_TE=0.92/0.7 msec, TR=5.2 msec, Flip angle=3deg, SENSE =1.5,1.5 in two directions. Breath-hold duration was 13.5 seconds, and a respiratory sensor was used to monitor compliance. The entire protocol lasted 15-20 minutes.

Data for each 2-echo mDIXON sequence was reconstructed on the MR system using the available standard single-peak spectral model of fat. Each sequence yielded four images per slice: water only, fat only, in-phase, out-phase. From the DICOM “water” and “fat” images, a pixel-wise “signal fat-fraction” parameter map was generated as follows using an offline post-processing tool. Fat percentage was calculated as (SI (Fat) x 100)/(SI(W+F)) where SI is the pixel signal intensity on the reconstructed fat and water images. Images were independently analyzed by a trained abdominal radiologist, documenting direct reading of percentage of fat in all eight hepatic segments using an ROI of 200-300 mm. Care was taken to select only representative regions, avoiding hepatic vasculature, biliary structures, or regions with artifacts.

## Results

### Bioprofiles accurately predict MRI-assessed liver fat infiltration

Alignment of model predictions with PDFF was assessed across 3 metrics: AUROC at 5%, 15%, and Spearman coefficient. In the validation set, Bioprofiles outperformed foundation model (CLMBR^26,27^), clinical (FSI^15^, HSI^16^), and technical benchmarks on all tasks (Table 2). Notably, CLMBR underperformed steatosis-specific clinical benchmarks at estimating PDFF.

**Table 2:** Model performance for predicting PDFF. All values are means (95% confidence interval) unless otherwise indicated.

|  | AUC (PDFF= 5%) | AUC (PDFF= 15%) | Spearman<br>Correlation vs.<br>PDFF |
| --- | --- | --- | --- |
| CLMBR (EHR-based Foundation Model) | 0.693 (0.681-0.705) | 0.716 (0.690-0.741) | 0.361 (0.345-0.377) |
| Framingham Steatosis Index (FSI) | 0.773 (0.744-0.802) | 0.751 (0.717-0.785) | 0.522 (0.472-0.566) |
| Hepatic Steatosis Index (HSI) | 0.793 (0.766-0.821) | 0.743 (0.709-0.777) | 0.542 (0.495-0.584) |
| Gradient Boosted (BMI/ALT/AST) | 0.800 (0.774-0.827) | 0.760 (0.727-0.794) | 0.569 (0.522-0.612) |
| Gradient Boosted (All Bioprofile input<br>features) | 0.827 (0.802-0.852) | 0.772 (0.739-0.805) | 0.604 (0.559-0.643) |
| Bioprofile | **0.851 (0.828-0.874)** | **0.801 (0.770-0.833)** | **0.650 (0.608-0.687)** |

To account for patterns of gaps in real-world data, we conducted sensitivity testing over data availability. We synthetically masked the fine-tuning cohort based on feature availability patterns learned from an external real-world EHR dataset to evaluate all models (Supplementary Table 2). Under these conditions, the Bioprofiles continued to outperform all benchmarks on all tasks.

To evaluate the generalizability of our models, we evaluated data from 5 clinical trials and hospital-based cohort studies where steatosis or MASLD status was assessed^32–36^. Bioprofiles continued to outperform clinical and technical benchmarks in all cases. (Table 3)

**Table 3:** External model validation on fatty liver status/biopsy (LBX)-confirmed steatosis. All values are means (95% confidence interval) unless otherwise indicated.

|  | Framingham<br>Steatosis Index<br>(FSI) | Hepatic Steatosis<br>Index (HSI) | Gradient Boosted<br>(All Bioprofile<br>input features) | Bioprofile |
| --- | --- | --- | --- | --- |
| Gifu Cohort<sup>34</sup> (AUROC vs ultrasound assessed MASLD status) (n=786) | 0.86 (0.83-0.89) | 0.84 (0.8-0.88) | 0.85 (0.82-0.89) | **0.88 (0.85-0.91)** |
| Kaohsiung Cohort<sup>35</sup> (AUROC vs ultrasound assessed MASLD severity) (n=407) | 0.69 (0.64-0.75) | 0.68 (0.63-0.74) | 0.68 (0.63-0.74) | **0.74 (0.69-0.78)** |
| Iksan Cohort<sup>36</sup> (Spearman R vs. CT-assessed disease severity) (n=913) | 0.25 (0.18-0.31) | 0.36 (0.29-0.42) | 0.35 (0.29-0.41) | **0.43 (0.37-0.49)** |

### Bioprofiles facilitate efficient subject nomination in simulation

To understand the impact of model improvements on study logistics, we conducted study simulations on each of our validation sets. For most model/task combinations, Portfolio Selection tended to outperform standard sampling, and Bioprofiles outperformed clinical benchmarks. However, for the targeted recruitment task, Bioprofiles alone were sufficient to achieve strong performance, likely due to the simpler desired distribution (Supplementary Tables 3, 4). However, on the even recruitment task, we observed a synergistic effect between the use of Bioprofile patient representations and the Portfolio Selection algorithm. To identify 12 subjects with PDFF > 10%, the Bioprofile required 19 screened subjects compared to 39 (HSI) and 43 (FSI). To identify 27 subjects in the even recruitment task, Bioprofiles required only 30 subjects screened, compared to 60 subjects (HSI) and 69 subjects (FSI) (Table 4). Notably, Bioprofile performance for the even recruitment task is close to ideal, with only three additional screenings projected to achieve recruitment goals. Study performance and variance for the even recruitment task was found to be a function of recruitment pool size (Supplementary Table 5).

**Table 4:**
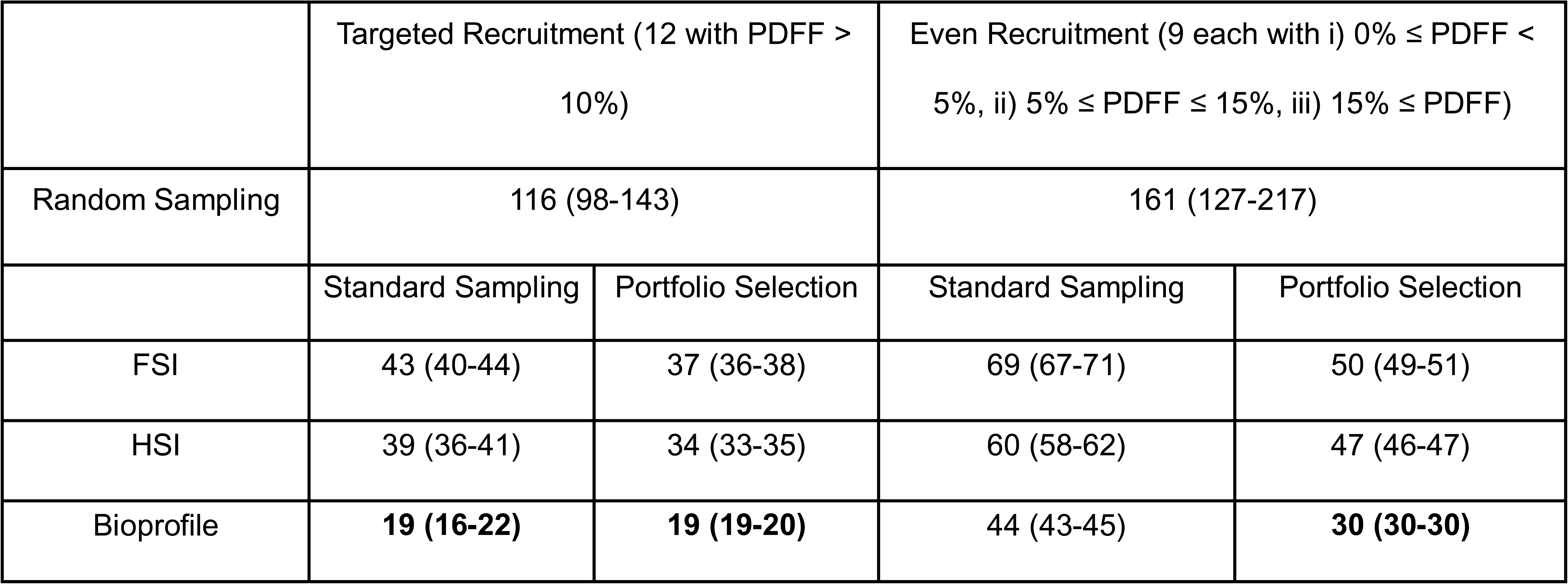
Simulated Recruitment Requirements for the fine-tuning set. All values are means (95% confidence interval) unless otherwise indicated.

### Bioprofiles successfully nominated of subjects for a human study

Using our Portfolio Selection algorithm, we identified a recruitment strategy that would enable us to embed the targeted recruitment task within the even recruitment task with minimal projected impact on total recruitment numbers (Supplementary Figure 1C). Recruitment was therefore conducted in two waves: first, the subpopulation collectively projected to have PDFF > 10% was recruited, followed by the subpopulations needed to obtain even recruitment. We deployed the student model over 45,484 individuals who met inclusion criteria in the VUMC CDW, and over 3 months of outreach, contacted 1,388 individuals through the MHAV platform. The targeted recruitment task was achieved after 20 subjects were screened, and the even recruitment task after 9 additional subjects, for 29 total screens. These values are in line with study simulations (Table 4) and result in substantially reduced numbers of excess screens relative to existing standards (Figure 1A, B). Next, we examined Bioprofile teacher model performance using deidentified pre-study and study visit data. We examined continuous measures of liver fat abundance, independent of bin definitions, comparing the Bioprofile predicted PDFF distribution to i) expected background and ii) actual observed distributions (Figure 1C). The predicted distribution was found to be significantly different from the background (KS test: p < 1E-3) but not significantly different to the distribution observed in enrolled study participants (KS test: p = 0.6). Finally, teacher Bioprofiles maintained strong subject level performance for PDFF among the recruited subjects (Figure 1D).

**Figure 1:**
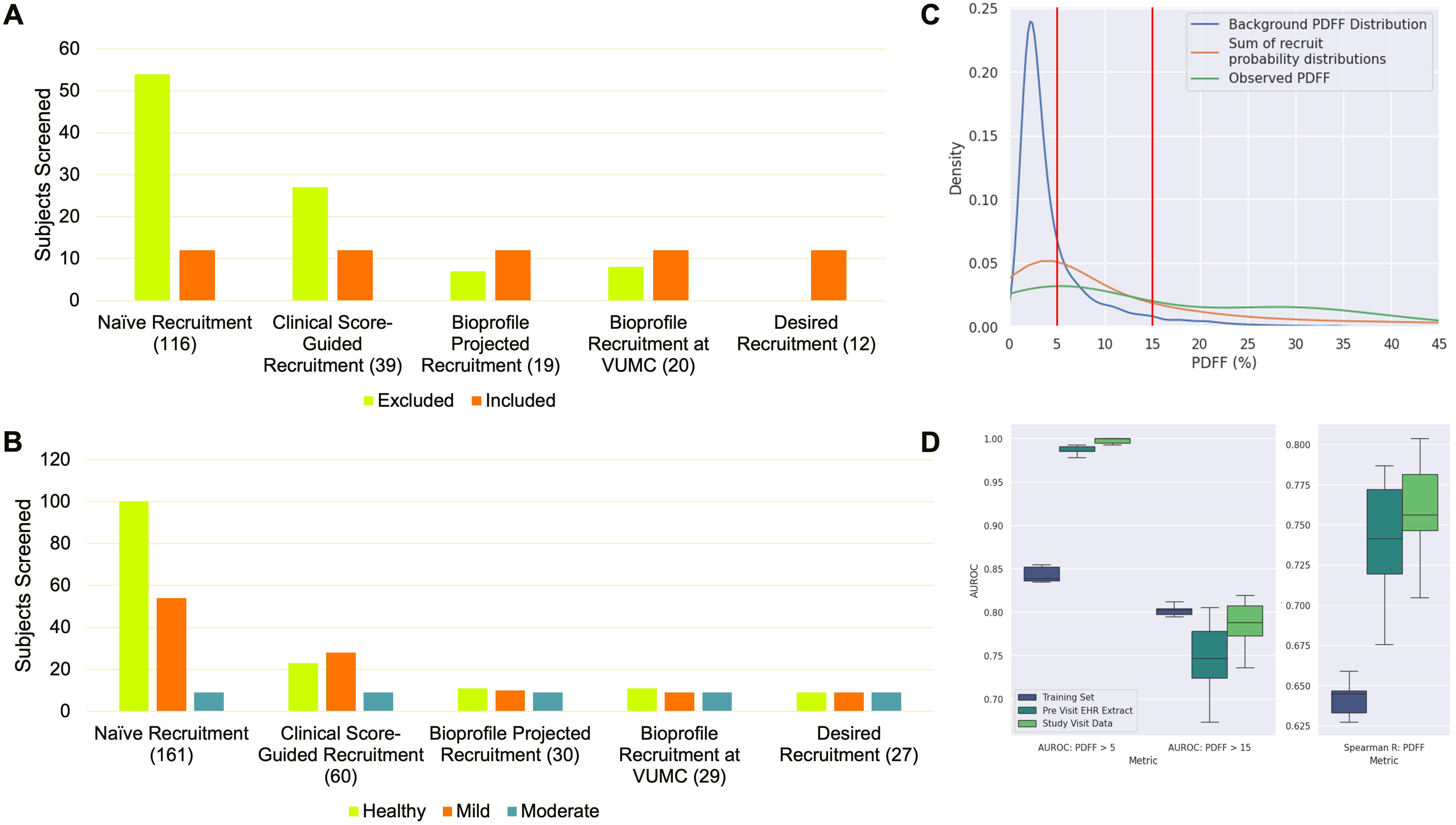
Results of VUMC Bioprofile Deployment Pilot: A, B) Bar charts of included/excluded subjects for targeted (A) and even (B) recruitment. Naïve sampling, clinical score and Bioprofile projected represent simulations from Table 4. C) Kernel density estimates for the background PDFF distribution (blue), Bioprofile predicted PDFF distribution (orange), and observed PDFF distribution (green). D) Subject level model performance on VUMC-recruited cohorts, stratified by input data type.

## Discussion

Our study demonstrates that AI-driven patient profiling can meaningfully surface subclinical disease signatures hidden in routine clinical data, providing a basis for streamlining clinical trial recruitment and enabling more proactive patient care. Using Bioprofiles, we efficiently nominated candidate participants for a prospective pilot study at VUMC, achieving high yield and strong agreement with imaging findings. By leveraging routine health record data pre-emptively, our approach enabled targeted invitations to patients who were most likely to qualify, thereby reducing screening failures. Simulation studies showed that the Bioprofile nomination pipeline halved the number of subjects that had to be screened to enroll the same number of eligible participants compared with conventional risk scores or untargeted approaches.

The relatively poor performance of an EHR-based foundation model on the PDFF prediction task highlights one of the intrinsic limitations of real-world data sources. Because the features captured in the EHR are typically generated through expressions of clinical judgement^37^, they do not provide complete representation of patient physiology. For diseases associated with substantial rates of underdiagnosis or clinical uncertainty, our approach of directly modeling accessible physiology is more likely to capture relevant disease states.

From a broader perspective, this work illustrates how a foundation model of cardiometabolic health can be translated into tangible recruitment benefits. While our study focused on trial recruitment, the underlying approach, estimating the results of less accessible tests using more accessible features, has much wider potential applications. Most immediately, Bioprofile patient representations have potential for providing probabilistic estimates for a wide series of cardiometabolic endpoints. Bioprofile-generated probability distributions can be easily incorporated into health economic or net-benefit calculations with the goal of optimizing care delivery and resource allocation. More directly, by mapping the overlap between Bioprofile output and existing clinical practice guidelines, Bioprofiles could act as a source of risk stratification or decision support.

Our findings should be interpreted in light of certain limitations. Our external validation focused on populations from Asia and trial participants. We are currently planning more extended validation in American community health contexts. Our VUMC pilot included 29 screens, which, while sufficient to demonstrate feasibility, is a small sample to fully establish the model’s performance characteristics. These subjects were also model-selected based on likelihood of meeting inclusion criteria and subject to response bias and so are not necessarily representative of the wider population. Our aggressive approach to feature mapping and curation was helpful in driving signal but was also highly labor intensive. Methods for interfacing or computing Bioprofiles from standard data models will be needed to enable wider-scale deployment. Finally, as currently configured, Bioprofiles are best suited to the highest prevalence indications among seemingly healthy primary care encounters. While we believe they can be extended to other indications, alternative datasets will be necessary to capture the full slate of “clinically accessible data” in other contexts.

In summary, this pilot highlights a promising future where AI-based physiological profiling becomes a routine tool for improving research efficiency and patient care. By leveraging routine health data to surface individuals with subclinical or unlabeled disease, Bioprofile modeling enabled precise and efficient recruitment in a MASLD-focused study, reducing the screening burden and improving alignment with inclusion criteria. This approach has the potential to accelerate drug development (through faster, more cost-effective trials) and to improve patient outcomes (through earlier detection and intervention). Beyond MASLD, applying Bioprofiles to identify subclinical cases could transform trial logistics and clinical care across a broad range of conditions by converting an invisible reservoir of patients into an actionable cohort for early identification, recruitment or discovery.

## Data Availability

Data Sharing Statement: The UK Biobank is available for use at https://www.ukbiobank.ac.uk by submitting a data request. The FLINT and NAFLD Adult databases are available for request at https://repository.niddk.nih.gov. The specific data from these studies used in this current work were used under license and so are not available. NHANES is freely available for use at https://www.cdc.gov/nchs/nhanes/index.html. The data from the Gifu, Kaohsiung, and Iksan cohorts are freely available at their respective journals. The Dandelion cohort was used under license and so is not available. The remaining data and analytic methods used in this study will remain the confidential property of the study sponsors. Data may be made available to other researchers upon request following permission, investigator support and following a signed data access agreement.

## Abbreviations

MASLD: metabolic dysfunction-associated steatotic liver disease
PDFF: proton density fat fraction.

## Acknowledgements

This research has been conducted using data from UK Biobank, a major biomedical database under application ID (76125).

