## Supplement for "Physiological foundation modeling for subclinical disease assessment: a prospective pilot"

**Supplementary Information:**

Embedding Development:

We utilized ReMasker, a masked autoencoder originally designed for tabular imputation, for the training of patient-level embeddings. Unlike standard autoencoders, ReMasker generates an embedding for each feature and utilizes transformers as part of its encoder and decoder blocks. As a result, there were multiple plausible intermediate embeddings that could be extracted from a trained ReMasker model. We evaluated all combinations of the following five elements: i) the CLS token from the encoder block, ii) the CLS token from the decoder block, iii) mean-pooling over feature tokens from the decoder, iv) max-pooling over feature tokens from the decoder, v) the imputed data (ReMasker’s natural output). In cross-validation, the combination of the i), iii) and v) had the strongest performance for predicting PDFF (Supplementary Figure 3). This embedding was used for all liver and non-liver-related tasks (Supplementary Table 1).

Sample-size calculation:

The prospective pilot was designed to demonstrate proof‑of‑concept rather than definitive clinical accuracy. The simulated target sample size of 30 was powered to detect a correlation coefficient as low as 0.49 (two-tailed α = 0.05, β = 0.2), compared to the observed correlation between Bioprofile predictions and PDFF of 0.646 (0.637-0.654). This target was therefore judged adequate for feasibility assessment within available resources.

Portfolio Selection Algorithm:

**Input:** ${P\in[0,1]}^{N\times T}$: Predicted bin-probabilities for $N$ patients across $T$ bins.

$d\in\mathbb{Z}_{\geq0}^{T}$: desired patient count per bin

$y\in\left\{ 1, \ldots,T \right\}^{N}$: true bin assignments for each patient

**Output:** $S \subseteq$ $\left\{ 1, \ldots,N \right\}$: selected patient indices that achieve the target count if feasible

$$r\leftarrow d$$

$$A\leftarrow\left\{ 1, \ldots,N \right\}, S\leftarrow\emptyset$$

**while** $\boldsymbol{(\exists}t\in\left\{ 1, \ldots,T \right\}$ such that $d_{t}>0$and $A\neq\emptyset$:

$B \leftarrow GETPORTFOLIO\left( P_{A}, r \right)$

**if** $B=\emptyset$ **then**

**break**

$S\leftarrow S\cup\left\{ A_{i}:i\in B \right\}$

**for** $t\leftarrow1$**to** $T$*:*

$c_{t} \leftarrow\left| \left\{ i\in B:y_{A_{i}}=t \right\} \right|$

$r_{t} \leftarrow max(0, r_{t}-c_{t})$

$A\leftarrow A \backslash\left\{ A_{i}:i\in B \right\}$

**return** $S$

GETPORTFOLIO algorithm

**Input:** ${P_{A}\in[0,1]}^{M\times T}$: Predicted bin-probabilities for $M$ patients across $T$ bins.

$r\in\mathbb{Z}_{\geq0}^{T}$: residual patient count per bin

**Output:** $B \subseteq$ $\left\{ 1, \ldots,M \right\}$: selected patient indices to be added this round

**Decision variables**: $x_{i}\in\left\{ 0,1 \right\}$ for $i=1,\ldots,M$, where $x_{i}$=1 if candidate $i$is selected

**Objective:** minimize $\sum_{i=1}^{M} x_{i}$ subject to **for** $t\leftarrow1$**to** $T$*:* $\sum_{i=1}^{M} P_{i,t}x_{i}\geq r_{t}$

Solve with PuLP, returning $x^{*}$ as the solution, else returning $\emptyset$

$$B\leftarrow\left\{ i\in\left\{ 1,\ldots,M \right\}:x_{i}^{*}=1 \right\}$$

**return** $B$

Study inclusion and exclusion criteria:

Subjects were included if they had preconsented to contact from researchers through the MHAV platform, had an email address on file, spoke English, were within the catchment area of VUMC (Davidson County, TN), and had been seen at VUMC for a primary care or outpatient encounter after January 1, 2022. Subjects were excluded if they had diagnoses of Severe pre-eclampsia, HELLP syndrome, Acute fatty liver of pregnancy, Maternal care for (suspected) damage to fetus from alcohol, not applicable or unspecified, Alcohol dependence in pregnancy, Diseases of the digestive system complicating pregnancy, childbirth and the puerperium, Newborn affected by maternal use of alcohol, Biliary atresia, Alagille Syndrome (Congenital malformation syndromes affecting multiple systems), Fetal alcohol syndrome (dysmorphic), Disorder of thorax, Toxic effect of alcohol, Finding of alcohol in blood, Starvation, High alcohol level in blood, Encounter for issue of repeat prescription, Long term drug therapy or Liver Transplant status. Subjects were excluded if they had prescriptions of atazanavir, darunavir (TMC114), fosamprenavir, indinavir, lopinavir/ritonavir, nelfinavir, ritonavir, saquinavir, tipranavir, Nucleoside/Nucleotide Reverse Transcriptase Inhibitors (NRTIs), abacavir, didanosine (ddI), emtricitabine (FTC), lamivudine (3TC), stavudine (d4T), tenofovir DF, zalcitabine (ddC), zidovudine (AZT), Non-Nucleoside Reverse Transcriptase Inhibitors (NNRTIs), delavirdine, efavirenz, Etravirine, nevirapine, enfuvirtide (T-20; fusion inhibitor), maraviroc (CCR5 antagonist), raltegravir (integrase inhibitor), Amiodarone, Tamoxifen, Methotrexate, Tacrolimus (FK506, Prograf), Cytoxan (cyclophosphamide), or Corticosteroids (systemic).

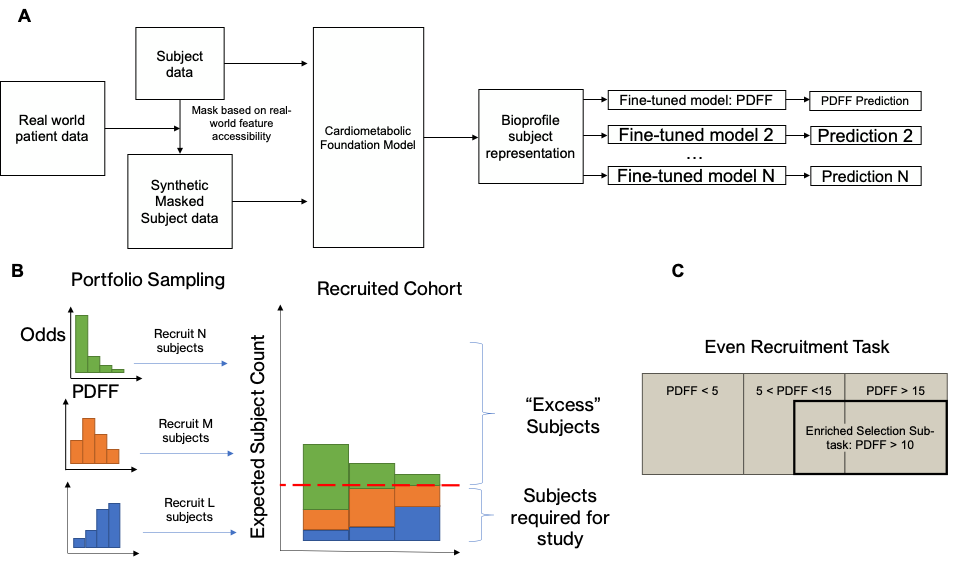

Supplementary Figure 1: Study Motivation and Methods Overview: A) Overview of Bioprofile training process. B) Overview of Portfolio Sampling algorithm. C) Embedding of Enriched Selection Task within Even Recruitment Task. The selection of the enriched PDFF > 10 sub-population was passed as an additional constraint to the Portfolio Sampler.

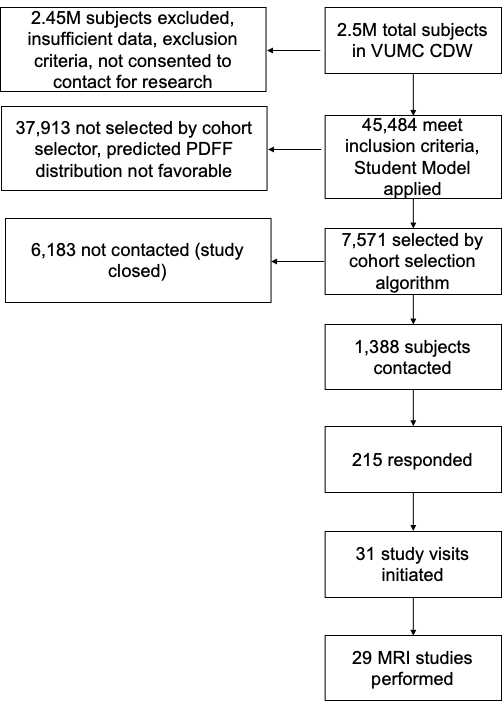

Supplementary Figure 2: CONSORT Diagram for subject recruitment at VUMC

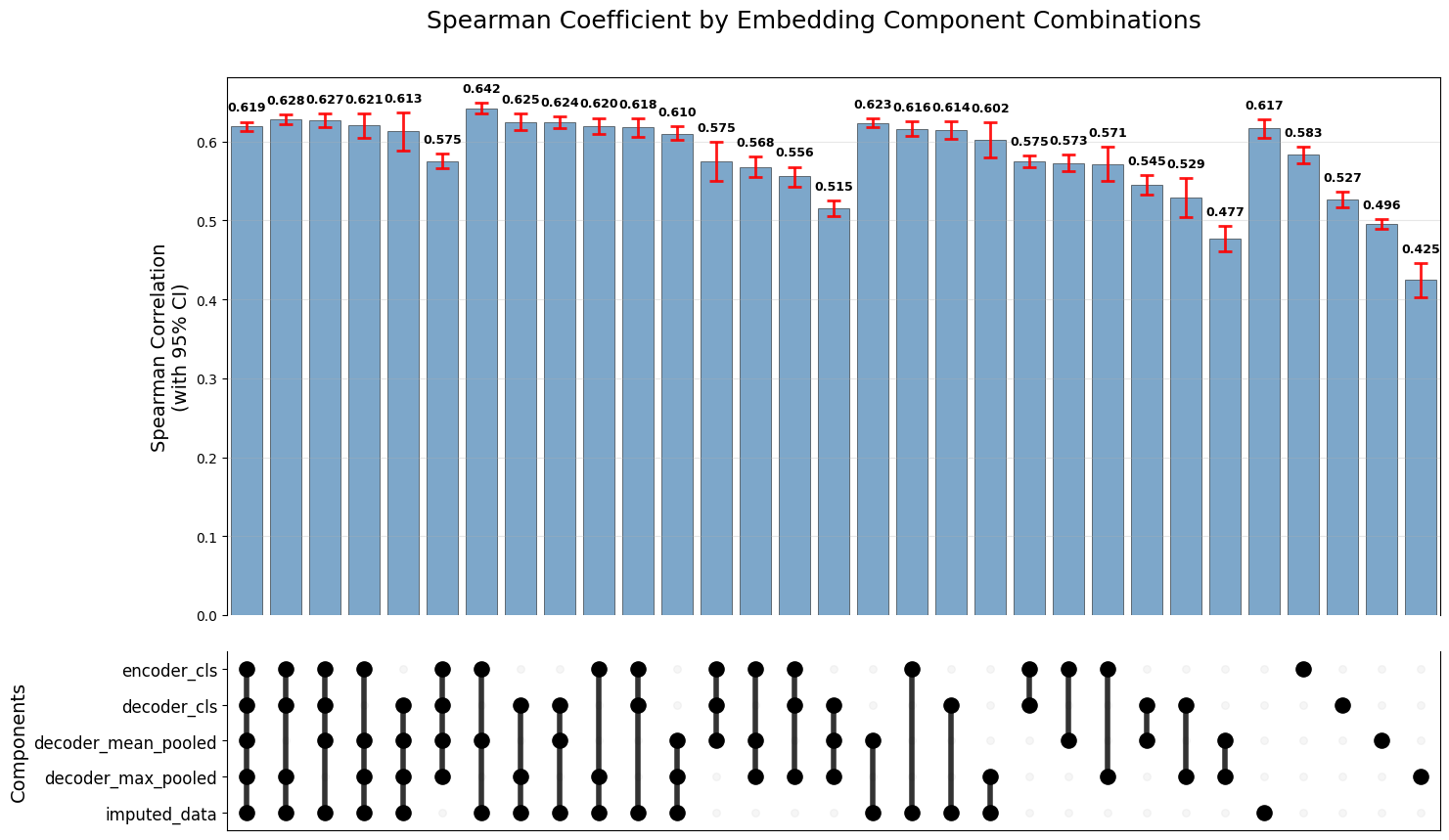

Supplementary Figure 3) Embedding composition determination for predicting PDFF using CARD regression. CIs are computed using cross-validation.

| **Endpoint** | **Bioprofile** | **Baseline Gradient Boosted Regressor** |
| --- | --- | --- |
| **Arterial Stiffness > 10 m/s (AUROC)** | **0.708 (0.0.688-0.729)** | 0.681 (0.664-0.698) |
| **FEV1/FVC > 0.7 (AUROC)** | **0.709 (0.691-0.728)** | 0.651 (0.631-0.671) |
| **Pancreas PDFF % (Spearman Coef)** | **0.608 (0.590-0.624)** | 0.577 (0.556-0.598) |
| **Liver cT1 > 800ms (AUROC)** | **0.817 (0.790-0.844)** | 0.805 (0.800-0.810) |
| **Grip Strength (mean Kg) (Spearman Coef)** | **0.809 (0.802-0.816)** | 0.767 (0.759-0.775) |

Supplementary Table 1: Model performance for predicting non-PDFF endpoints. All values are means (95% confidence interval) unless otherwise indicated.

|  |  | **AUC (Liver Fat = 5%)** | **AUC (Liver Fat = 15%)** | **Spearman Correlation vs. Liver Fat** |
| --- | --- | --- | --- | --- |
| **CLMBR** | Longitudinal Only | 0.601 (0.578-0.624) | 0.598 (0.596-0.610) | 0.222 (0.207-237) |
|  | RW Missingness Mask | 0.691 (0.679-0.703) | 0.696 (0.671-0.721) | 0.358 (0.342-0.375) |
|  | Full Input Data | 0.693 (0.681-0.705) | 0.716 (0.690-0.741) | 0.361 (0.345-0.377) |
| **FSI** | RW Missingness Mask | 0.758 (0.729-0.788) | 0.733 (0.698-0.767) | 0.489 (0.441-0.533) |
|  | Full Input Data | 0.773 (0.744-0.802) | 0.751 (0.717-0.785) | 0.522 (0.472-0.566) |
| **HSI** | RW Missingness Mask | 0.785 (0.757-0.813) | 0.727 (0.692-0.762) | 0.521 (0.471-0.566) |
|  | Full Input Data | 0.793 (0.766-0.821) | 0.743 (0.709-0.777) | 0.542 (0.495-0.584) |
| **Gradient Boosted Baseline** | RW Missingness Mask | 0.807 (0.781-0.833) | 0.741 (0.707-0.775) | 0.553 (0.504-0.594) |
|  | Full Input Data | 0.827 (0.802-0.852) | 0.772 (0.739-0.805) | 0.604 (0.559-0.643) |
| **Bioprofile** | RW Missingness Mask | 0.817 (0.792-0.843) | 0.763 (0.730-0.796) | 0.583 (0.535-0.625) |
|  | Full Input Data | **0.851 (0.828-0.874)** | **0.801 (0.770-0.833)** | **0.650 (0.608-0.687)** |

Supplementary Table 2: Model performance for predicting PDFF using the resampled VUMC validation set under real-world missingness conditions. All values are means (95% confidence interval) unless otherwise indicated. CLMBR was evaluated under separate conditions: one where data from UK Biobank study visits and longitudinal primary data were both available, but the RW mask was applied (RW Missingness Mask), and one where the longitudinal primary care data only was available, but no mask was applied (Longitudinal Only).

| Random Sampling | 116 (98-143) | |
| --- | --- | --- |
|  | Standard Sampling | Portfolio Selection |
| FSI | 43 (40-44) | 37 (36-38) |
| HSI | 39 (36-41) | 34 (33-35) |
| Gradient Boosted (BMI/ALT/AST) | 29 (27-31) | 25 (24-26) |
| Gradient Boosted (All Bioprofile input features) | 28 (27-32) | 27 (26-28) |
| Bioprofile (Student) | N/A | 23 (20-27) |
| Bioprofile (Teacher) | 19 (16-22) | **19 (19-20)** |
| Ideal Performance | 12 | |

Supplementary Table 3: Simulated Recruitment Requirements for targeted recruitment task for all benchmarks. All values are means (95% confidence interval) unless otherwise indicated.

| Random Sampling | 161 (127-217) | |
| --- | --- | --- |
|  | Standard Sampling | Portfolio Selection |
| FSI | 69 (55-57) | 50 (49-51) |
| HSI | 60 (58-62) | 47 (46-47) |
| Gradient Boosted (BMI/ALT/AST) | 54 (53-55) | 57 (54-60) |
| Gradient Boosted (All Bioprofile input features) | 47 (46-48) | 46 (43-49) |
| Bioprofile (Student) | N/A | 31 (31-32) |
| Bioprofile (Teacher) | 44 (43-45) | **30 (30-30)** |
| Ideal Performance | 27 | |

Supplementary Table 4: Simulated Recruitment Requirements for even recruitment task for all benchmarks. All values are means (95% confidence interval) unless otherwise indicated.

| Recruitment Pool Size | Portfolio Selection + Bioprofile, Subjects needed to screen |
| --- | --- |
| 300 | 55.8 (40-71.6) |
| 500 | 39 (35-43.2) |
| 1000 | 35.9 (30.8-41) |
| 2500 | 36.6 (29.9-43.3) |
| 5000 | 35.4 (29-41.8) |
| 7500 | 32.7 (30.3-35.1) |
| 10000 | 30 (30-30) |

Supplementary Table 5: Portfolio Selection/Bioprofile Study performance for Even Recruitment Task as a function of Recruitment Pool Size. All values are means (95% confidence interval) unless otherwise indicated.

Supplementary Table 6: Hyperparameters used for ReMasker training

| learning_rate | 0.0065 |
| --- | --- |
| embed_dim | 96 |
| decoder_depth | 4 |
| num_heads | 12 |
| mlp_ratio | 4.64 |
| weight_decay | 0.00001 |
| batch_size | 2560 |
| mask_ratio | 0.68 |

Supplementary Table 7: Hyperparameters used for CARD training

| learning_rate | 0.0048 |
| --- | --- |
| n_hidden_mean_model | 64, 256 |
| n_hidden_guidance_model | 128, 128, 32 |
| beta_schedule | linear |
| beta_start | 8.8e-5 |
| beta_end | 0.285 |
